# Laying the Foundation for *iCANmeditate*: A mixed methods study protocol for understanding patient and oncologist perspectives on meditation

**DOI:** 10.1101/2023.08.21.23294379

**Authors:** Yasmin Lalani, Alexandra Godinho, Kirsten Ellison, Krutika Joshi, Aisling Curtin Wach, Punam Rana, Pete Wegier

## Abstract

**Background:** People with cancer experience heightened levels of stress and anxiety, including psychological or physical. In recent years, digitally delivered complimentary therapies, such as meditation, have gained attention in cancer research and advocacy communities for improving quality of life. However, most digital meditation resources are commercially available and are not tailored to the unique needs of cancer patients (addressing fears of recurrence). As such, this study lays the foundation to co-design a publicly available digital meditation program called *iCANmeditate* that contains cancer-specific meditation content.

**Aims:** To understand: (1) cancer patients’ perceptions and practices of meditation, as well as their needs in addressing the stress that accompanies their cancer diagnosis and (2) current knowledge of meditation and prescribing trends amongst oncologists in Canada.

**Methods and analysis:** A mixed-methods design comprised of online patient and oncologist surveys and a series of patient focus groups will be used. Survey data analysis will use multivariate logistic regressions to examine predictors of: (1) interest in using a meditation app among patients and (2) prescribing meditation among oncologists. Patient focus groups will gather insights about the contexts of daily living where meditation would be most beneficial for people with cancer; this data will be analyzed thematically.

**Discussion:** The results of this study will inform iterative co-design workshops with cancer patients to build the digital meditation program *iCANmeditate*; focus group results will be used to develop vignettes or “personas” that will supply the initial stimulus material for the iterative co-design workshops. Once the program has been finalized in partnership with cancer patient participants, a usability and pilot study will follow to test the functionality and efficacy of the tool. Results from the oncologist survey will form the basis of knowledge mobilization efforts to facilitate clinical buy-in and awareness of the benefits of meditation to cancer patients.

## Introduction

Most people with cancer experience heightened levels of stress, which can continue post-recovery for many.(1, 2) This stress can take several forms including psychological and physical stress, as well as stress that accompanies patients’ fears of uncertainty about the future.(3) Such stressors have been associated with changes in immune system function, negative mental health outcomes, and reduced quality of life.(4) Moreover, some of the core symptoms that have been identified as requiring improved management strategies among cancer patients go hand-in-hand with stress such as: anxiety, depression, fatigue, pain, and insomnia.(5) Although several medications can assist with the management of these symptoms, cancer patients have increasingly become interested in using complementary therapies due to their perceived safety, potential for reducing side effects, and improving overall well-being.(6, 7)

One complementary therapy that has shown positive outcomes for patients to manage their cancer-related symptoms is meditation.(8, 9) Meditation is a mind-body practice that involves focusing one’s attention on the present to achieve mental clarity. As a whole, meditation can refer to several different techniques (e.g., body-centered, mindful, or contemplative) and relies on different bodily mechanisms to focus one’s attention (e.g., breath, sounds, visualizations).(10) With its roots in Eastern spiritual traditions, it has gained widespread appeal in healthcare and clinical practice guidelines as an evidence-based therapy for improving mental health (i.e., anxiety, depression, psychological distress), physical health (e.g., sleep, fatigue, pain, feeling sick), and overall quality of life in those affected by cancer.(11, 12)

The uptake of meditation in cancer populations is also rising, marked by surveys demonstrating over one third of patients use it as a form of complementary therapy,(6) and via the growing number of studies examining its efficacy across different types and stages of cancer.(13–17) Moreover, a general shift towards offering meditation as a digital resource has been observed within the last decade, as compared to traditional in-person sessions. Many scholars argue that online or smartphone-based meditation apps offer cancer patients greater flexibility and access to care, by removing some of the reported challenges of attending face-to-face classes such as: lack of nearby services, competing interests for time (e.g., appointments, caregiver availability, childcare), and committing to travel and lengthy sessions when distressed and fatigued.(18–20) In addition, recent findings have also shown a preference among cancer patients for using smartphone applications to learn about support care services, particularly among younger and non-white patients thereby making digital meditation a potentially viable support service.(21)

Commercially available digital meditation tools have shown promise in cancer patients, with initial efficacy trials demonstrating these tools to be comparable to in-person sessions in terms of acceptability, feasibility, and positive outcomes for cancer populations.(22–24) However, despite the numerous publicly available digital meditation tools on the market (e.g., Calm^TM^, Headspace^TM^) that have shown promise in cancer patients, few are tailored to the specific needs and preferences of people affected by cancer.(14, 20, 25) Indeed, findings from a recent study suggest that cancer patients would prefer to use digital meditation tools that are optimized to their needs and preferences.(8, 26) In particular, Huberty and colleagues revealed that cancer patients who used the commercially available Calm^TM^ app for meditation, have expressed modifications to Calm^TM^ to contain information about how to center on positive feelings, how to manage negative feelings associated with their cancer journey, and content that promotes movement-based activities.(26)

To address this gap and provide a publicly available solution, we propose to co-design a digital meditation tool called *iCANmeditate* that is tailored to the specific needs of cancer patients and leverages an existing hospital-based digital platform. We will be conducting this study with the guidance of one of the authors, Dr. Punam Rana who is a medical oncologist and certified meditation teacher (https://drpunamrana.com/). However, to accomplish this our team must first understand: (1) patient perceptions and practices of meditation as well as the needs and concerns of cancer patients’ experiences with stress and coping with their diagnosis and (2) current knowledge of meditation and prescribing trends amongst oncologists. To date, only one study has examined the knowledge and perceptions of meditation among cancer patients in preparation for designing a tool.(27) Overall, this study found that while the benefits of meditation were generally understood by cancer patients, one of the main barriers to engaging with meditation was a lack of perceived knowledge. In addition, these scholars found that experience with meditation use seemed to vary by gender, age, and education level, however, interest in a digital resource was highly correlated with reporting a higher level of stress.(27) Although these findings are important for understanding the characteristics of cancer patients who may be more likely to use and benefit from a digital meditation tool, this study was limited to melanoma patients and is not generalizable to all cancer types and stages.

## Aim of the study

We propose to conduct a survey and in-depth focus groups on patient knowledge, prior use, and attitudes towards meditation as it relates to their experiences of stress and coping with their diagnosis. In addition, we also propose to survey Canadian oncologists on their current knowledge and attitudes towards meditation, as no study to date has been conducted in this area to our knowledge. The survey will ask oncologists about their awareness of different digital meditation resources and clinical practice guidelines, as well as current practices in prescribing meditation to their patients. As this study is exploratory in nature, no a priori hypotheses were made.

## Methods

### Participants

#### Patients

Outpatients attending the Humber River Health Cancer Clinic will be invited to take part in a survey about well-being, including strategies for coping with their stress and anxiety. Eligibility criteria will consist of being 18 years of age or older, currently receiving treatment for cancer and English-speaking; for focus group participants they will have indicated on the survey that they would be interested to engage in focus groups. Individuals will not be excluded based on type or stage of cancer, however those with cognitive impairments, psychiatric illness, or are too distressed to participate as identified by clinicians will be excluded. The following types of cancer will be excluded from the sample as they are not currently treated at the clinic: head, neck, sarcoma, skin, melanoma, gynecological cancers.

#### Oncologists

Members of the Canadian Association of Medical Oncologists (CAMO) will be invited to take part in a brief online survey about their perspectives on prescribing meditation to improve quality of life in cancer patients. CAMO is comprised of over 350 members that include: medical oncologists, residents, fellow, and associates. Eligibility criteria will consist of being 18 years of age or older, having completed their fellowship, and practicing medical oncology.

### Study Design and Procedures

#### Patient Survey and Focus Groups

Eligible participants will be identified through Humber River Health’s Cancer Clinic outpatient treatment appointment lists, and clinical eligibility will be determined in consultation with a clinical nurse coordinator. Recruitment will take place in the cancer clinic treatment area, with research staff systematically approaching all patients in treatment bays who have not been identified by nursing staff as being on airborne precautions, significantly distressed, or cognitively unable to provide consent. Patients will be invited to participate in a survey about cancer patient’s well-being, including experiences of stress, anxiety, and quality of life. To ensure that patients in the clinic understand research activities are not a part of standard clinical care, posters will be placed in the cancer clinic waiting room. To reduce self-selection biases (i.e., recruiting only participants who are interested in meditation), and capture a wide range of patients’ perceptions and barriers towards meditation, participants will be blinded to the purpose of the survey. Consent forms and recruitment scripts will not include any mention of meditation. Interested potential participants will be directed to an online consent form by research staff on their individual integrated bedside terminal (i.e., tablet); this is a confidential and private tablet available to each patient receiving treatment in the cancer clinic. Informed consent will include an optional consent to be contacted for future studies and anonymous data sharing (i.e., open science). To ensure consent is informed, potential participants will be provided with the opportunity to ask questions and express concerns to research staff, with teach-back strategies being used as needed.(28) Those who complete the consent form will be redirected to an online survey. Upon survey completion participants will be debriefed about the true aims of the study (i.e., to understand patient perspectives and experience with meditation in preparation of developing a digital meditation tool) and will be provided with a patient resource about meditation. If participants feel some discomfort in reflecting about their wellbeing and quality of life even after they complete the survey, there is a distress centre phone number indicated on the consent form. In addition, all participants who complete the survey will be provided with a $5 physical gift card as a token of appreciation.

Participants who indicate on the survey that they would be interested in using a digital meditation resource and participating in online focus groups to help develop a cancer-specific meditation program will be considered for participation in virtual focus groups. Participant characteristics and experience with meditation will be used to guide the selection of participants; a maximum of 15 participants will be recruited for two independent focus groups. This will be a purposive, homogeneous sample of approximately six to eight participants per focus group, with a cancer diagnosis, and some experience with meditation. Focus groups with an overarching homogeneous composition can be desirable to create a comfortable environment for self-expression amongst participants who have a shared experience.(29, 30) Participants who are selected for participation will be emailed a consent form outlining the study in greater detail. To ensure consent is informed, participants will be encouraged to discuss questions and concerns with research staff via email or phone calls prior to consenting to participate; again teach-back strategies will be used when appropriate.(28) Those who complete the consent form will be invited to attend up to four online focus groups. All participants who engage in focus groups will be compensated with the equivalent of $25/hour in the form of an electronic gift card (maximum compensation of $200 if participants attend 4 focus group sessions). All focus group sessions will be video, and audio recorded as well as transcribed verbatim. The video files will be permanently deleted, and the audio files will be kept for analysis. Risks of participating in focus groups may be momentarily discomfort of talking about their cancer experiences or hearing others’ experiences; if participants still feel distress after the focus group sessions, they may refer to the Distress Centres of Greater Toronto phone number indicated on the consent form as a way to seek professional help. All identifying information from audio transcripts will be removed.

#### Oncologist Survey

Medical oncologists who are members of CAMO will be invited to complete a survey about their perspectives and knowledge of meditation for improving quality of life in cancer patients via an email survey blast. To promote recruitment efforts, up to two reminders will be sent 1-week post-initial survey email blast, and first reminder, respectively. Those who are interested in completing the survey will be directed to an electronic consent form via a link contained in the invitation email. Potential participants who complete the electronic consent will be automatically directed to the survey on their browser. Upon completion of the survey, oncologists will be provided with a meditation resource that they can view and download. Participants will be compensated with a $5 electronic gift card.

#### Meditation Resources

All meditation resources in this study provide a plain-language overview of meditation and draw on existing meditation resources provided by the Canadian Cancer Society, the National Center for Complementary and Integrative Health, and the Memorial Sloan-Kettering Cancer Centre. All materials have been reviewed for accuracy by the clinical subject matter lead for meditation.

#### Patient Meditation Resource

Following the completion of the patient survey, participants will be offered a tri-fold brochure that provides a plain-language overview of what meditation is and how it has been shown to benefit cancer patients. The brochure will also provide them with a link and QR code to Dr. Punam Rana’s website, where they will have access to additional resources and information.

#### Oncologist Meditation Resource

A one-page document will also be made available at the end of the survey to oncologists who express interest in learning more about meditation (i.e., respond “*yes”* to survey item asking about interest in meditation resources). This electronic pdf will provide oncologists with a plain-language overview of what meditation is and how it has been shown to benefit cancer patients. The document will also provide them with references to current clinical guidelines and a list of relevant online resources and recommended patient resources.

### Measures and Qualitative Methods

#### Patient Survey Measures

The survey will assess a variety of demographic characteristics that have been associated with meditation practice such as: age, sex at birth, education level, employment, and religion. In addition, clinical characteristics such as cancer diagnosis details (i.e., type, stage, date of diagnosis) and mental health outcomes will also be assessed.(31, 32)

##### Mindfulness

Everyday mindfulness will be measured using the Cognitive and Affective Mindfulness Scale-Revised (CAMS-R), which measures the following aspects of mindfulness: present-focused attention, awareness, and nonjudgmental acceptance of thoughts and emotions.(33) This tool has been shown to be suitable for use in meditating as well as non-meditating samples, and is commonly used in cancer patient samples to assess individual differences in dispositional mindfulness.(27) This study will use the brief 10-item version of the tool, where scores range from 10 to 40,with higher scores representing greater self-reported mindfulness.

##### Experience With Meditation

A series of questions have been developed by the team, in consultation with a meditation expert (Dr. Punam Rana), to assess patients’ practice of meditation. In particular, participants will be asked about: type of meditation practiced (e.g., mindful, transcendental); resources used (e.g., apps, books, websites); tools to assist in meditating (e.g., incense, mat, bells); duration and frequency of practice (months/years, daily/weekly/monthly); and changes in meditation since their cancer diagnosis (decision to meditate/frequency). In addition, participants will also be asked about current interest in learning more about meditation, continuing their current practice, and the extent to which they feel their practice of meditation is helpful.

##### Barriers to Meditation

Perceived barriers for meditating among cancer patients who currently meditate, versus those who do not practice meditation will be assessed using the Determinants of Meditation Practice Inventory – Revised (DMPI-R).(34) This briefer version of the full DMPI,(35) is a psychometrically validated 12-item self-report tool that measures 4 subscales of barriers to meditating: low perceived benefit, perceived inadequate knowledge, perceived pragmatic barriers, and perceived socio-cultural conflict. Each subscale is comprised of 2 - 4 items that are scored on a 5-point Likert-type scale, with higher scores indicating a higher perception of that perceived group of barriers.

##### Knowledge of Meditation

Patient knowledge of meditation will be assessed using a series of questions that gauge general perceptions of meditation. A set of 10 items have been developed by Russell et al.(27) to assess knowledge of meditation. Survey respondents are asked to rate their agreement with 10 statements (i.e., seven facts and three misconceptions that are reverse scored), using the response options 1 “Agree”, 0 “Don’t know” or -1 “Disagree. Overall, higher total scores indicate greater knowledge of meditation.

#### Focus Groups

The purpose of the focus groups will be to explore participants’ (1) experiences, perceptions, and preferences regarding meditation and (2) how and under what contexts their practice of meditation has (or has not) helped the cope with their cancer diagnosis. The focus group guide is structured based on focus group question route methodology; general questions will be asked first followed by more specific key questions directly related to the research topic so that participants are given ample time to “warm up” and feel comfortable with one another.(36, 37)

The moderator of the focus groups will be a member of the research team with qualitative research expertise. Additionally, a notetaker and technical support person will be present with their cameras off. The notetaker will capture non-verbal interactions amongst participants and any other data which would add to a fuller picture of the focus group event. The technical support person will be available to troubleshoot any issues with the Zoom platform that participants may have, as well as monitor the chat function.

To capture the majority of themes, it is preferable to build in more than one focus group.(38) Thus, after the first focus group session, we will invite participants to return for a second, third, or fourth focus group session until data saturation has been reached. Depending on availability, we will invite the same participants to all focus groups; however, if there is a demand for focus group participation, we may conduct further focus groups with different participants from the survey participant pool.

#### Oncologist Survey Measures

The survey will assess demographic characteristics such as participant age and sex as well as professional practice characteristics including main place of work, years and province of practice, and area of oncology specialty. To determine current practices around recommending alterative and complementary therapies for alleviating stress and anxiety among cancer patients, participants will be asked to identify any therapies they currently recommend from a pre-specified list of the most common forms (e.g., music therapy, meditation, acupuncture); an open-ended user option will also be included to capture therapy forms not listed.

##### Meditation Recommendation Practices

To assess the proportion of survey completers who specifically recommend meditation, an additional yes/no question will be asked to those who do not select meditation from the aforementioned list (i.e., “Have you *ever* recommended meditation to your patients?”). In addition, for those who respond that they do or have recommended meditation, further questions will assess the forms of meditation that have been recommended (i.e., types of meditation vs. delivery format), and how the recommendation was made. In order to better understand how oncologists recommend meditation to their patients, an item was developed to assess the physicians’ level of engagement in providing information and resources about meditation. This item will ask “Which statement best describes how you recommend meditation to your patients?” and provide the following response options: (a) I suggest meditating to help with stress and anxiety, (b) I recommend meditation to my patients and provide them with resources to help them learn and understand how to meditate, and (c) I explain the benefits of meditation to my patients and provide them with resources to help them learn and understand how to meditate.

##### Knowledge of Meditation

The survey will also aim to understand oncologists’ attitudes and knowledge of meditation, by asking the same series of questions outlined for patient participants; see above.(27)

##### Awareness of Practice Guidelines Endorsing Meditation

The last component of the survey aims to understand oncologists’ general awareness for guidelines that currently recommend meditation to cancer patients. In 2018, the American Society for Clinical Oncology (ASCO), endorsed guidelines put forth by the Society for Integrative Oncology (SIO), recommending music therapy, meditation, stress management, and yoga for anxiety and stress reduction among breast cancer patients. Survey respondents will be presented with a partial screenshot of the practice guidelines and items will be asked to assess (1) knowledge/awareness for the guidelines, (2) if their recommendations of integrative therapies changed upon release of the guidelines, and if so what therapies they started recommending to their patients, and (3) for those without awareness for the guidelines, if becoming aware of them via the survey would influence their recommendation of integrative therapies moving forward as well as which specific therapies they will begin recommending to patients.

## Data Management

The electronic platform, Qualtrics, used to collect consent and participant data for both the patient and oncologist online surveys ensures research participants’ privacy and confidentiality, and does not share data with third parties. Survey responses on Qualtrics are protected by Qualtrics’ Web Application Firewalls and their detection system that monitors for unauthorized users. Data will not be linked from other sources and will only be used for purposes outlined in the consent documentation. Patient and oncologist digital survey data will be stored in a secure computer file and will be deleted upon the completion of the study. A Master Linking Log that contains patients’ Study IDs and personal identifying information will be kept as a password protected file, accessible to the research team during the recruitment process only. Focus group audio files will be sent to a transcription service using secure file transfer; transcripts and fieldnotes will be anonymized and stored both in a locked office (if they are printed as paper copies) as well as digitally as password protected digital files. All study data (i.e., survey data/focus group transcripts) will be stored securely for at least five years on Humber River Health’s encrypted cloud storage, after which the de-identified data will be deposited into Humber River Health’s Institutional Repository for a period of 10 years. The de-identified data will not be sold and will only be accessible to the study team.

## Data analysis and sample size calculations

### Patient Survey Data Analysis and Sample Calculation

The primary analysis will be a prediction of who is interested in using a meditation app, applying multivariate Logistic Regression with two-sided p<0.05 as critical value. The covariates include age, anxiety level, perceived stress, and perceived mindfulness while the factors include sex and whether the patient uses alternate methods of stress management. In secondary analyses, we will use bivariate statistical analyses to identify relationships, interactions, or differences between the covariates/factors using T-tests and Spearman correlations. Based on the number of covariates/factors (k=6), and a lower estimate of the proportion of the positive outcome variable (40%; v=0.4), the minimum number of cases required is calculated by the formula (N=10k/v), as determined by Peduzzi, et al.(39) This results in a calculated sample size of 150 for a meaningful logistic regression analysis.

### Patient focus group data analysis

Analysis of focus groups will require a dynamic, non-linear, and iterative engagement with the following data items: transcripts, fieldnotes, selective listening of audio, photos of any activities captured during the recording of the online focus group such as: Poll Everywhere, Zoom Whiteboard notes in the chat etc. The moderator team will meet to debrief immediately after the focus group to discuss particular moments in the session that are noteworthy or that may require special attention during analysis.

All data forms listed above will be uploaded into NVivo 12 qualitative analysis software program as the majority of the analysis steps will be conducted in this program. To ensure that analysis is trustworthy, focus group data will be analyzed by the qualitative lead in the research team. The approach to analysis will be iterative and performed in sequence; when each focus group is complete, data analysis will begin prior to the next scheduled focus group. This will allow time for theoretical sampling to take place for the subsequent focus group, in the event that a particular theme or topic requires refining. As each focus group is completed, data will be analyzed using the constant comparison method which includes (1) coding the data into small units and attaching a descriptor or ‘code’ to each unit, (2), grouping the codes into larger categories (axial coding), and (3) developing broader themes from larger categories.(40, 41)

The lead analyst will work with the note-taker to discuss codes and themes that are under construction during the analysis process. The purpose of these discussions is twofold: (1) to achieve consensus regarding the meanings and definitions of codes as well as resulting themes to resolving and (2) to incorporate any non-verbal communicative data from the recording as well as other dynamic moments of interaction captured via notes rather than solely the transcript. In doing so, we will eliminate as much as possible, the potential misleading interpretations of participants’ voices analyzed as words on the transcript only.

### Oncologist survey data analysis and sample calculation

The primary analysis will be a prediction of who prescribes meditation using multivariate Logistic Regression with two-sided *p*<0.05 as critical value. The covariates include age while the factors include sex and whether alternate therapies are recommended. In secondary analyses, we will use multiple regression to determine relationships between the covariates/factors. We will also test for interactions, particularly between whether alternate therapies are recommended and who prescribes meditation. Based on the number of covariates/factors (k=3), and a lower estimate of the proportion of the positive outcome variable (20%; v=0.2), the minimum number of cases required is calculated by the formula (N=10k/v), where 10 is the minimum number of ‘events per variable’ as determined by Peduzzi, et al.(39) This results in a calculated sample size of 150 for a meaningful logistic regression analysis. As a result, all oncologists belonging to the Canadian Institution of Medical Oncologists (CAMO) will be given the survey.

## Discussion

### Significance of the proposed study

The study outlined in this protocol aligns with recent research in healthcare interventions and delivery that empowers patients to engage with digital solutions to manage their symptoms and enhance their wellbeing.(42, 43) Cancer patients in particular experience stress, pain, and decreased quality of life as a consequence of their diagnosis; some patients would therefore benefit greatly from a digital tool for symptom management. Thus, this study responds to the most recent call for online meditation programs for cancer to be tailor-made for cancer patients as opposed to standard meditation digital applications.(8, 26)

This study protocol serves as the first step to address this gap; it outlines the rationale and procedures for understanding cancer patients’ perceptions and practices of meditation as well as their experiences of how meditation may have benefited them. In tandem with the quantitative and qualitative patient data, the survey results of practicing oncologists in Canada will aid in our understanding of their prescribing practices and state of meditation knowledge. Taken together, the patient and oncologist datasets will prepare the infrastructure needed for the next phase of the broader research goal—to co-design and pilot test an online meditation program called *iCANmeditate*.

The co-design process will be informed by vignettes generated from patient focus group data. Vignettes are fictional representations or “personas” of individuals’ experiences of the topic under study and have practical applications in healthcare in co-design or participatory health research.(44) Vignettes will be presented to cancer patient participants during the development phase of this larger research program to: (1) develop content and a prototype of the online meditation tool on the hospital’s platform and (2) to test the usability of the meditation program. Co-design workshops will be run iteratively with feedback incorporated by participants until *iCANmeditate* is complete. A pilot study will follow the development of *iCANmeditate* to measure the tool’s efficacy.

### Strengths and Limitations

One key strength of this study is our engagement with oncologists in Canada; there is a paucity of research that demonstrates the level of knowledge and prescribing behaviours of meditation as a complimentary integrative therapy from oncologists despite the fact that meditation is included in some clinical guidelines.(11, 45) Consequently, a long-term goal for the meditation program is to convey the value of meditation by involving oncologists so that meditation does not continue to be a “fringe” therapy with little clinical endorsement.

Possible limitations of this foundational study can be pinpointed. One limitation is that the patient population drawn from the hospital study site may not be representative of patients with cancer at a national or provincial level. For instance, the patient survey excludes participants whose command of English is weak and as such, survey results will only reveal data from an English-speaking group. Secondly, the hospital study site does not treat all types of cancer and thus, a comprehensive understanding of patients’ knowledge of meditation will exclude patients with head, neck, sarcoma, gynecological, skin and melanoma cancers. In a similar vein, the target population for the patient survey is patients who are actively in treatment and not patients who were recently diagnosed. Our rationale for excluding this group was to avoid adding an emotional burden to patients who may still be coming to terms with their cancer diagnosis. Finally, our patient sample will not include individuals who may identify as “survivors”; the concept of survivorship in cancer is an emerging field of study and consequently, outside the scope of this investigation of cancer patients currently in treatment.

In conclusion, this protocol outlines the research activities required to move to our next phase of research where continuous engagement with cancer patients during the co-design phase will produce a unique and meaningful meditation program that incorporates the specific needs of people with cancer. This protocol has been approved by Veritas Independent Review Board, Tracking Number: 2023-3319-15429-4.

## Data Availability

No datasets were generated or analyzed during the current study. De-identified research data will be made publicly available when the study is completed and published.

## Acknowledgements

We would like to thank the Cancer Care Clinic at Humber River Health for their continuing support of this research program.

## Notes

### Competing Interest Statement

The authors have declared no competing interest.

### Funding Statement

The authors received no specific funding for this work.

### Author Declarations

This protocol has been approved by Veritas Independent Review Board, Tracking Number: 2023-3319-15429-4.

## References

1. Bates GE, Mostel JL, Hesdorffer M. Cancer-related anxiety. JAMA oncology. 2017;3(7):1007-.

2. Cillessen L, Johannsen M, Speckens AEM, Zachariae R. Mindfulness-based interventions for psychological and physical health outcomes in cancer patients and survivors: A systematic review and meta-analysis of randomized controlled trials. Psychooncology. 2019;28(12):2257–69.

3. Børøsund E, Mirkovic J, Clark MM, Ehlers SL, Andrykowski MA, Bergland A, et al. A stress management app intervention for cancer survivors: design, development, and usability testing. JMIR formative research. 2018;2(2):e9954.

4. Antoni MH, Dhabhar FS. The impact of psychosocial stress and stress management on immune responses in patients with cancer. Cancer. 2019;125(9):1417–31.

5. Reeve BB, Mitchell SA, Dueck AC, Basch E, Cella D, Reilly CM, et al. Recommended Patient-Reported Core Set of Symptoms to Measure in Adult Cancer Treatment Trials. J Natl Cancer Inst. 2014;106(7).

6. Huebner J, Prott FJ, Micke O, Muecke R, Senf B, Dennert G, et al. Online survey of cancer patients on complementary and alternative medicine. Oncology research and treatment. 2014;37(6):304–8.

7. Tangkiatkumjai M, Boardman H, Walker D-M. Potential factors that influence usage of complementary and alternative medicine worldwide: a systematic review. BMC Complementary Medicine and Therapies. 2020;20(1):1–15.

8. Klainin-Yobas P, Hounsri K, Chng WJ, Ang NKE, Goh Y-SS. Preventing psychological symptoms among cancer survivors through a digital mindfulness psychoeducation program: Protocol of a randomized controlled trial. medRxiv. 2023:2023.01. 05.23284230.

9. Tacón AM. Meditation as a complementary therapy in cancer. Family and Community Health. 2003;26(1):64–73.

10. Matko K, Sedlmeier P. What is meditation? Proposing an empirically derived classification system. Frontiers in psychology. 2019;10:2276.

11. Greenlee H, DuPont-Reyes MJ, Balneaves LG, Carlson LE, Cohen MR, Deng G, et al. Clinical practice guidelines on the evidence-based use of integrative therapies during and after breast cancer treatment. CA: a cancer journal for clinicians. 2017;67(3):194–232.

12. Horowitz S. Health benefits of meditation: What the newest research shows. Alternative and Complementary Therapies. 2010;16(4):223–8.

13. Huberty J, Eckert R, Larkey L, Joeman L, Mesa R. Experiences of Using a Consumer-Based Mobile Meditation App to Improve Fatigue in Myeloproliferative Patients: Qualitative Study. JMIR Cancer. 2019;5(2):e14292.

14. Huberty J, Puzia M, Eckert R, Larkey L. Cancer patients’ and survivors’ perceptions of the calm app: cross-sectional descriptive study. JMIR cancer. 2020;6(1):e16926.

15. Puzia ME, Huberty J, Eckert R, Larkey L, Mesa R. Associations between global mental health and response to an app-based meditation intervention in myeloproliferative neoplasm patients. Integrative cancer therapies. 2020;19:1534735420927780.

16. Russell L, Ugalde A, Livingston PM, Orellana L, Milne D, Krishnasamy M, et al. A pilot randomised controlled trial of an online mindfulness-based program for people diagnosed with melanoma. Supportive Care in Cancer. 2019;27(7):2735–46.

17. Swainston J, Derakshan N. Reduced anxiety following mindfulness and adaptive working memory training in women with breast cancer. Mindfulness. 2021;12(8):1928–39.

18. Gowin K, Millstine D, Kosiorek HE, Langlais B, Huberty J, Eckert R. The SIMM study: survey of integrative medicine in myeloproliferative neoplasms. Haematologica. 2017;4:102–92.

19. Kubo A, Altschuler A, Kurtovich E, Hendlish S, Laurent CA, Kolevska T, et al. A pilot mobile-based mindfulness intervention for cancer patients and their informal caregivers. Mindfulness. 2018;9:1885–94.

20. Zernicke KA, Campbell TS, Speca M, Ruff KM, Flowers S, Tamagawa R, et al. The eCALM trial: eTherapy for cancer applying mindfulness. exploratory analyses of the associations between online mindfulness-based cancer recovery participation and changes in mood, stress symptoms, mindfulness, posttraumatic growth, and spirituality. Mindfulness. 2016;7:1071–81.

21. Raghunathan NJ, Korenstein D, Li QS, Tonorezos ES, Mao JJ. Determinants of mobile technology use and smartphone application interest in cancer patients. Cancer medicine. 2018;7(11):5812–9.

22. Atreya CE, Kubo A, Borno HT, Rosenthal B, Campanella M, Rettger JP, et al. Being present: a single-arm feasibility study of audio-based mindfulness meditation for colorectal cancer patients and caregivers. PLoS One. 2018;13(7):e0199423.

23. Huberty J, Bhuiyan N, Puzia M, Joeman L, Larkey L, Mesa R. Meditation Mobile App Developed for Patients With and Survivors of Cancer: Feasibility Randomized Controlled Trial. JMIR cancer. 2022;8(4):e39228.

24. Mikolasek M, Witt CM, Barth J. Effects and implementation of a mindfulness and relaxation app for patients with cancer: Mixed methods feasibility study. JMIR cancer. 2021;7(1):e16785.

25. Kubo A, Kurtovich E, McGinnis M, Aghaee S, Altschuler A, Quesenberry Jr C, et al. A randomized controlled trial of mHealth mindfulness intervention for cancer patients and informal cancer caregivers: a feasibility study within an integrated health care delivery system. Integrative cancer therapies. 2019;18:1534735419850634.

26. Huberty J, Bhuiyan N, Neher T, Joeman L, Mesa R, Larkey L. Leveraging a Consumer-Based Product to Develop a Cancer-Specific Mobile Meditation App: Prototype Development Study. JMIR Formative Research. 2022;6(1):e32458.

27. Russell L, Orellana L, Ugalde A, Milne D, Krishnasamy M, Chambers R, et al. Exploring Knowledge, Attitudes, and Practice Associated With Meditation Among Patients With Melanoma. Integrative Cancer Therapies. 2018;17(2):237–47.

28. Kripalani S, Bengtzen R, Henderson LE, Jacobson TA. Clinical research in low-literacy populations: using teach-back to assess comprehension of informed consent and privacy information. IRB: Ethics & Human Research. 2008;30(2):13–9.

29. Robinson J. Using focus groups. Handbook of qualitative research in education: Edward Elgar Publishing; 2020. p. 338–48.

30. Woźniak W. Homogeneity of Focus Groups as a Pathway to Successful Research Findings? Przegląd Socjologii Jakościowej. 2014;10(1):6–23.

31. Cohen S. Perceived stress in a probability sample of the United States. 1988.

32. Spitzer RL, Kroenke K, Williams JB, Löwe B. A brief measure for assessing generalized anxiety disorder: the GAD-7. Archives of internal medicine. 2006;166(10):1092–7.

33. Feldman G, Hayes A, Kumar S, Greeson J, Laurenceau J-P. Mindfulness and emotion regulation: The development and initial validation of the Cognitive and Affective Mindfulness Scale-Revised (CAMS-R). Journal of psychopathology and Behavioral Assessment. 2007;29:177–90.

34. Hunt CA, Hoffman MA, Mohr JJ, Williams A-l. Assessing perceived barriers to meditation: The determinants of meditation practice inventory-revised (DMPI-R). Mindfulness. 2020;11:1139–49.

35. Williams A-l, Dixon J, McCorkle R, Van Ness PH. Determinants of meditation practice inventory: development, content validation, and initial psychometric testing. Alternative Therapies in Health and Medicine. 2011;17(5):16.

36. Gill P, Stewart K, Treasure E, Chadwick B. Methods of data collection in qualitative research: interviews and focus groups. British dental journal. 2008;204(6):291–5.

37. Plummer-D’Amato P. Focus group methodology Part 1: Considerations for design. International Journal of Therapy and Rehabilitation. 2008;15(2):69–73.

38. Guest G, Namey E, McKenna K. How many focus groups are enough? Building an evidence base for nonprobability sample sizes. Field methods. 2017;29(1):3–22.

39. Peduzzi P, Concato J, Kemper E, Holford TR, Feinstein AR. A simulation study of the number of events per variable in logistic regression analysis. Journal of clinical epidemiology. 1996;49(12):1373–9.

40. Krueger RA. Focus groups: A practical guide for applied research: Sage publications; 2014.

41. Onwuegbuzie AJ, Dickinson WB, Leech NL, Zoran AG. A qualitative framework for collecting and analyzing data in focus group research. International journal of qualitative methods. 2009;8(3):1–21.

42. Martínez-Pérez B, De La Torre-Díez I, López-Coronado M. Mobile health applications for the most prevalent conditions by the World Health Organization: review and analysis. Journal of medical Internet research. 2013;15(6):e120.

43. Zheng C, Chen X, Weng L, Guo L, Xu H, Lin M, et al. Benefits of Mobile Apps for Cancer Pain Management: Systematic Review. JMIR mHealth and uHealth. 2020;8(1):e17055.

44. Hughes R, Huby M. The application of vignettes in social and nursing research. Journal of advanced nursing. 2002;37(4):382–6.

45. Mao JJ, Ismaila N, Bao T, Barton D, Ben-Arye E, Garland EL, et al. Integrative medicine for pain management in oncology: society for integrative oncology–ASCO guideline. Journal of Clinical Oncology. 2022;40(34):3998–4024.

